# Heterogeneity of comprehensive clinical phenotype and longitudinal adaptive function and correlation with computational predictions of severity of missense genotypes in KIF1A-associated neurological disorder

**DOI:** 10.1101/2024.02.29.24303377

**Authors:** Khemika K. Sudnawa, Wenxing Li, Sean Calamia, Cara H. Kanner, Jennifer M. Bain, Aliaa H. Abdelhakim, Alexa Geltzeiler, Caroline M. Mebane, Frank A. Provenzano, Tristan T. Sands, Robert J. Fee, Jacqueline Montes, Yufeng Shen, Wendy K. Chung

**Affiliations:** Department of Pediatrics, Boston Children’s Hospital, Harvard Medical School, Boston, MA, USA; Department of Pediatrics, Phramongkutklao Hospital and Phramongkutklao College of Medicine, Bangkok, Thailand; Department of Systems Biology, Columbia University Irving Medical Center, New York, New York, USA; Department of Pediatrics, Columbia University, New York, New York, USA; Department of Rehabilitation and Regenerative Medicine, Columbia University Irving Medical Center, New York, New York, USA; Departments of Neurology and Pediatrics, Columbia University Irving Medical Center, New York, New York, USA; Harkness Eye Institute, Department of Ophthalmology, Columbia University Irving Medical Center, New York, NY, USA; Virginia Tech Carilion School of Medicine; Taub Institute for Research on Alzheimer’s Disease and the Aging Brain, Department of Neurology, Columbia University Medical Center, New York, New York, USA; Departments of Neurology and Pediatrics, Vagelos College of Physicians and Surgeons, Columbia University, New York, New York, USA; Department of Neurology, Columbia University Vagelos College of Physicians and Surgeons and New York-Presbyterian Hospital, New York, New York, USA; Department of Biomedical Informatics, Columbia University Irving Medical Center, New York, New York, USA

**Keywords:** KIF1A, hypotonia, spasticity, seizures, ataxia, optic nerve atrophy, cerebellar atrophy, cognitive impairment, adaptive function, ESM

## Abstract

**Purpose:** Pathogenic variants in Kinesin Family Member 1A (*KIF1A*) are associated with *KIF1A*-associated neurological disorder (KAND). We report the clinical phenotypes and correlate genotypes of individuals with KAND.

**Methods:** Medical history and adaptive function were assessed longitudinally. In-person evaluations included neurological, motor, ophthalmologic and cognitive assessments.

**Results:** We collected online data on 177 individuals. Fifty-seven individuals were also assessed in-person. Most individuals had *de novo* heterozygous missense likely pathogenic/pathogenic *KIF1A* variants. The most common characteristics were hypotonia, spasticity, ataxia, seizures, optic nerve atrophy, cerebellar atrophy, and cognitive impairment. Mean Vineland Adaptive Behavior Composite score (VABS-ABC) was low (*M*=62.9, *SD*=19.1). The mean change in VABS-ABC over time was -3.1 (*SD*=7.3). The decline in VABS-ABC was associated with the age at first assessment and abnormal electroencephalogram/seizure. There was a positive correlation between Evolutionary Scale Model (ESM) score for the variants and final VABS-ABC (*p*=0.003). Abnormal electroencephalogram/seizure, neuroimaging result, and ESM explain 34% of the variance in final VABS-ABC (*p*<0.001).

**Conclusion:** In-person assessment confirmed caregiver report and identified additional visual deficits. Adaptive function declined over time consistent with both the neurodevelopmental and neurodegenerative nature of the condition. Using ESM score assists in predicting phenotype across a wide range of unique variants.

## Introduction

Pathogenic variants in Kinesin Family Member 1A (*KIF1A*) (OMIM#601255) are associated with *KIF1A*-Associated Neurological Disorder (KAND). Pathogenic *KIF1A* variants are most commonly heterozygous *de novo* variants associated with a wide range of neurodevelopmental and neurodegenerative phenotypes. Inherited heterozygous and biallelic variant are also rarely observed^1^. Heterozygous variants contribute to heterodimeric proteins, leading to a dominant negative effect^2,3^.

The *KIF1A* gene encodes an adenosine triphosphate-dependent microtubule motor protein that is a member of the kinesin-3 family. KIF1A contains a motor, coiled-coil with a Forkhead associated, and a Pleckstrin-Homology (PH) domain. The motor domain moves along microtubules, and the PH domain binds the cargo. Cargo includes mature synaptic vesicles, their precursors, dense core vesicles, and Tropomyosin receptor kinase A-containing vesicles^4^. KIF1A also plays role in neuronal stem cell proliferation and brain development^5^.

In this study, we expanded our previously reported cohort^1^ to 177 individuals with pathogenic/likely pathogenic *KIF1A* variants and characterized them longitudinally. We also included in-person clinical evaluations for 57 individuals. We aimed to extend our understanding of the natural history of KAND over the life course.

## Materials and Methods

This study was approved by the Columbia University Institutional Review Board, and informed consent was obtained from all individuals or their guardians. The study included individuals with *KIF1A* pathogenic or likely pathogenic variants according to the American College of Medical Genetics classification guidelines^6^. All individuals previously had exome sequencing, genome sequencing, or panel gene testing, and many also had a chromosome microarray. Clinical genetic test reports were reviewed by a geneticist, and individuals with dual genetic diagnoses affecting the nervous system were excluded.

### Online clinical data collection

Medical data were collected by an interview with a physician or genetic counselor via phone. Follow-up clinical data were collected via online questionnaires completed by caregivers with an interval of at least 12 months. The clinical data were validated with medical records.

Adaptive function was assessed by using the Vineland Adaptive Behavior Scales 3rd Edition (VABS)^7^ comprehensive parent/caregiver form via the online Pearson’s Q-global™ platform. Longitudinal VABS follow-ups were administered at least 12 months apart (Supplemental Table 1).

Clinical electroencephalograms (EEG) and magnetic resonance imaging (MRI) were collected and reviewed by a child neurologist specializing in epilepsy and a neuroradiologist, respectively (Supplemental Table 1).

### In-person clinical assessments

In-person clinical assessments were conducted at Columbia University or at the KIF1A.ORG family meeting. The assessment included neurological and functional assessment by a child neurologist, motor assessment by a physical therapist, ophthalmic assessment by a genetic ophthalmologist, and cognitive assessment by a neuropsychologist (Supplemental Table 1).

Individual’s function was assessed using the Gross Motor Function Classification System (GMFCS)^8^, Manual Ability Classification System (MACS)^9^, Communication Function Classification System (CFCS)^10^, and Eating and Drinking Ability Classification System (EDACS)^11^. The Pediatric Feeding Oral Intake Scale (pFIOS) was also used to assess functional oral intake of food and liquids in patients with oropharyngeal dysphagia^12^.

In-person motor assessment: Gross motor function was assessed using the Gross Motor Functional Measure-88 (GMFM)^13,14^ which converted to GMFM-66 scores. Those who were able also completed upper extremity assessments using the Box and Blocks Test (BBT)^15^ and 9-Hole Peg Test (9HPT)^16^. Additionally, individuals who were ambulatory completed the 10-meter Walk Run test (10MWR)^17^, 6-Minute Walk Test (6MWT)^18^, Timed Up and Go test (TUG)^19,20^ and quantitative gait analysis on a Zeno instrumented walkway (Prokinetics, Havertown, PA)^21^.

In-person cognitive assessment: the Differential Ability Scales Second Edition (DAS-II) and the Wechsler Adult Intelligence Scale Fourth Edition (WAISLIV) were performed for individuals within age ranges of 2.5-17.9 and 18.0 years and older, respectively. Individuals less than 2.5 years of age, unable to perform the assessment, or not English proficient were excluded.

### Correlations between clinical and genetic factors with VABS scores

#### Data organization

Categorical variables were converted into quantitative variables to facilitate statistical analysis. Baseline EEG results and seizure status were combined into one variable, and individuals with an abnormal EEG or seizure were categorized as affected, and those with normal or unknown EEG and no seizure were categorized as unaffected for electrophysiology. The final Vineland ABC standard scores (VABS-ABC) were the most recently performed VABS for those with multiple measurements.

#### Evolutionary Scale Model (ESM) score of *KIF1A* variants

We obtained ESM-1b log likelihood ratio scores, which were predictions of genetic effect of missense variants by protein deep learning language models^22^, from https://huggingface.co/spaces/ntranoslab/esm_variants for all missense variants (Supplemental Table 2). A smaller value in ESM score indicates more damaging consequence to the protein function.

## Statistical analysis

Statistical analyses were conducted using IBM SPSS (version 28) and R statistical software version 4.2.2. Categorical variables are reported as frequency and percentages. Continuous variables with normal distributions are reported as the mean (*M*) with standard deviation (*SD*); variables with non-normal distributions are reported as median with interquartile range (*IQR*) or range. One-way analysis of variance with post hoc comparisons using the Bonferroni test were used to compare means of two or more independent groups. Pairwise correlation of clinical phenotypes using the “cor” function and the Spearman correlation coefficients for all paired variables were calculated. Principal components analysis (PCA) of clinical phenotypes was using the “factoextra” package in R, and the variables containing more than 20% missing values were removed. The correlation matrix of all input variables used the “ggcorrplot” function, the biplot combined with the squared cosine (cos2) used the “fviz_pca_var” function, and the contribution of all variables on each principal component used the “fviz_cos2” function. Differences in VABS-ABC and change of VABS-ABC between the two groups were using Student’s t-test. Univariate linear regression was used to analyze the correlation of phenotypes and VABS-ABC. Multivariate linear regression was used to analyze the correlation of major phenotypes and VABS-ABC with adjusted covariables. A *p* value ≤0.05 was considered as significant.

## Results

One hundred seventy-seven individuals were enrolled with a median age of 6.9 years (range 0.5-58 years). (Supplemental Figure 1). One hundred-seven individuals completed the first follow-up, and sixty-three individuals completed the second follow-up (Supplemental Figure 2). Fifty-seven individuals were assessed in-person with the median age of 8.3 years (range 1.7-59.5).

### Genetic variants

Almost all individuals had heterozygous likely pathogenic or pathogenic *KIF1A* heterozygous variants (Figure 1). One individual had biallelic variants. Most variants were missense variants (94.9%), 5 splice variants, 3 inframe indel variants, 1 frameshift variant, and 1 entire coding sequence deletion. Most variants were in the motor domain (97.7%). Variants were demonstrated to be *de novo* in 63.8%, inherited in 7.4%, and of unknown inheritance due to lack of parental testing in 28.8%. Five variants were inherited from parent (2 siblings) and seven variants were inherited from mosaic parents (1 sibling). Parents were not included in this cohort.

**Figure 1:**
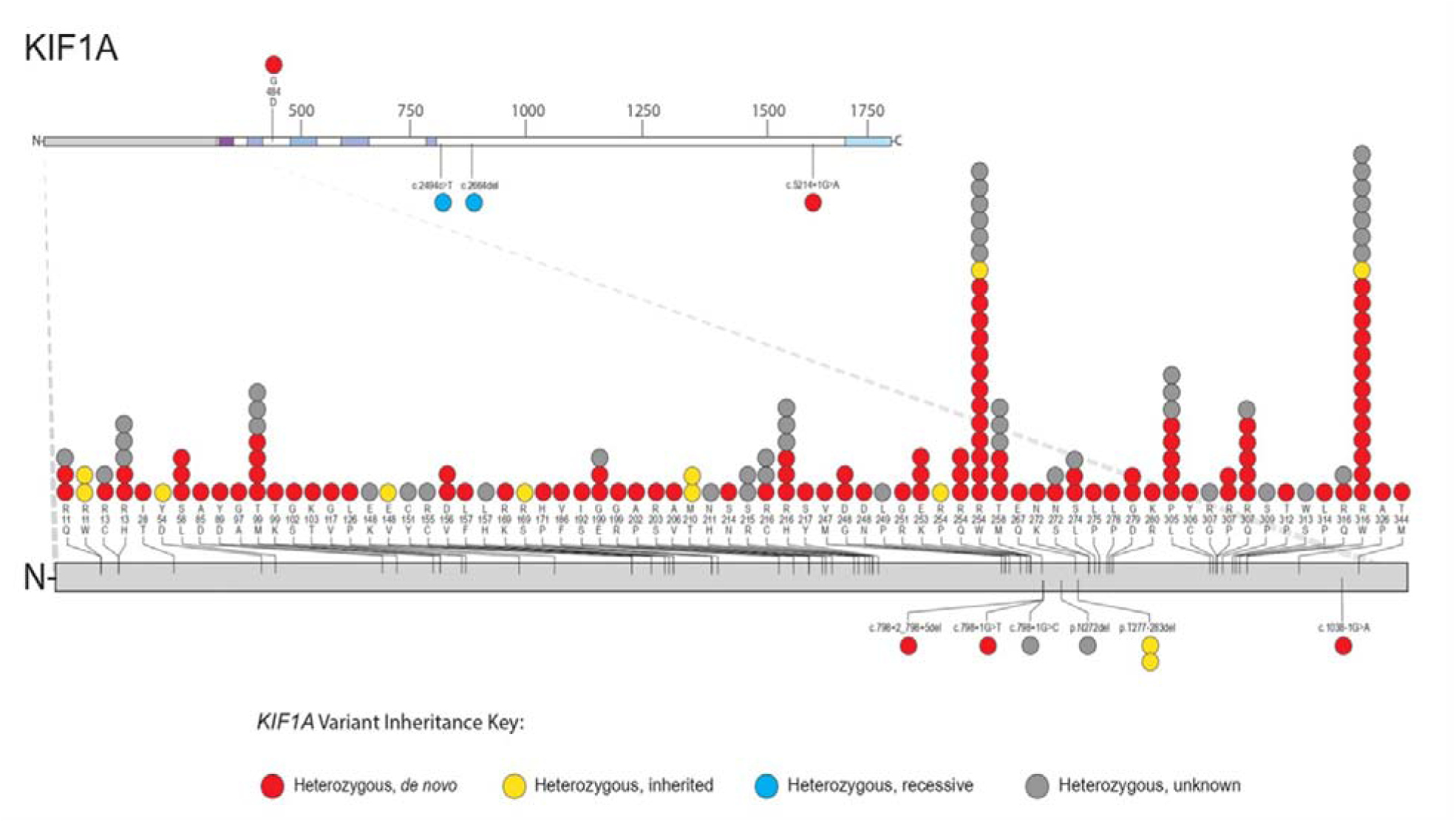
*KIF1A* likely pathogenic or pathogenic variants Each circle represents a single unrelated person with pathogenic/likely pathogenic *KIF1A* variants in this cohort that is color coded by inheritance. One individual with entire coding sequence deletion is not mapped in this figure. The top portion of the figure is the full-length protein, and the bottom portion of the figure is an enlargement of the motor domain. Missense variants are shown above the gene, other types of variants (splice variants, inframe indel, frameshift) are shown below the gene.

## Clinical characteristics in individuals with *KIF1A* variants

The clinical symptoms by caregiver report were aggregated across all ages (Table 1 and Supplemental Table 3). The median age of the individuals at last follow up was 9.1 years (range 0.6-59.4). For individuals <20 years old, the mean BMI z-score was 0.3 (*SD*=1.8, range -8.0 to 2.9). The mean BMI in adults was 25.7 kg/m^2^ (*SD*=5.0). The mean height z-score was -1.1 (*SD*=1.8).

**Table 1:** Clinical phenotypes in individuals with *KIF1A* Variants.

| Clinical characteristics | Percentage |
| --- | --- |
| Caregiver report |  |
| Gender (n=177): Male | 58.4 |
| Neurological problems (n=175) |  |
| - Hypotonic | 86.3 |
| - Hypertonic | 79.4 |
| - Excessively clumsy or uncoordinated | 61.1 |
| - Movement abnormalities | 60.6 |
| - Seizures | 41.1 |
| - Peripheral neuropathy | 32.0 |
| - Microcephaly | 15.4 |
| Seizure type (n=72) |  |
| - Absence seizure | 70.8 |
| - Generalized tonic-clonic seizure | 44.4 |
| - Spasms seizure | 34.7 |
| - Atonic seizure | 33.3 |
| - Complex partial seizure | 20.8 |
| - Simple partial seizure | 18.1 |
| - Intractable seizure | 9.7 |
| Visual problems (n=172) |  |
| - Optic nerve atrophy | 52.9 |
| - Cortical visual impairment | 26.7 |
| - Strabismus | 31.4 |
| - Nystagmus | 22.1 |
| - Myopia | 30.2 |
| - Astigmatism | 21.5 |
| - Hyperopia | 16.3 |
| - Depth perception problems | 22.1 |
| - Amblyopia | 14.0 |
| - Cataracts | 8.7 |
| Neurological examination (n=49) |  |
| Muscle atrophy |  |
| - Both legs atrophy | 51 |
| - Diffuse atrophy | 8.2 |
| Tone |  |
| Hypotonia |  |
| - Axial hypotonia | 73.5 |
| - Upper extremity hypotonia | 51 |
| - Lower extremities hypotonia | 16.3 |
| Spasticity |  |
| - Bilateral MAS1 | 59.2 |
| - Bilateral MAS2 | 6.1 |
| - Asymmetric lower extremities spasticity (MAS1/MAS2) | 4.1 |
| Weakness | 59.2 |
| Hyperkinetic movement disorder | 20.4 |
| - Stereotypies | 12.2 |
| - Tics | 4.1 |
| - Dystonia | 2 |
| - Self-injurious movements | 2 |
| Coordination abnormalities | 24.5 |
| Musculoskeletal |  |
| - Pes planus | 55.1 |
| - Contractures | 10.2 |
| - Scoliosis | 10.2 |
| - Cavovarus abnormality | 4.1 |
| - Hip dysplasia | 4.1 |
| - Kyphosis | 2 |
| - Navicular drop | 2 |
| Note from neurological examination<br>- Several demonstrated mottling of the skin with purple and blue discoloration of the extremities and several had cold feet.<br>- Contractures were present in the toes, ankles, hips and fingers.<br>MAS=Modified Ashworth Scale |  |

### Neurological symptoms

By caregiver report, neurological symptoms included hypotonia (86%), hypertonia (79%), and difficulty with coordination (61%) with a median age of onset at 1 (*IQR*=1.1), 2 (*IQR=*3.2), and 1.6 (*IQR*=2.8) years, respectively. Movement abnormalities were reported in 61% with a median age of onset at 1.6 years (IQR=2.8), including ataxia (28%), tremors (22%), and dystonia (15%). Peripheral neuropathy was noted in 32%, and microcephaly in 15%. Forty-one percent of individuals had a history of seizures with a median age of diagnosis at 3.3 years (*IQR*=4). The most common seizure type was absence seizures (71%). Thirty-eight percent of individuals with seizures reported >100 lifetime episodes of seizure and 31% experienced seizures daily. Many different antiseizure medications were used (Supplemental Table 4); and 49.2% of individuals with seizures had seizures in the past 6 months.

#### Electroencephalogram

By caregiver report of 121 individuals with EEGs performed, 64 (53%) reported abnormal results. There were 18 individuals with EEG tracings available for review. Median age at the time of the recordings was 4 years (IQR 4.4). Of these 18 individuals, 15 (83%) had abnormal studies due to either slowing of the EEG background (13 individuals) and/or epileptiform abnormalities (12 individuals). Epileptiform discharges were focal in 10 individuals and generalized in 2 individuals.

#### Neuroimaging

By caregiver report, neuroimaging abnormalities were noted in 55%, with previously normal neuroimaging in 21%. The median age of abnormal neuroimaging was 2 years (*IQR*=2.2). The most common brain abnormalities were cerebellar atrophy (74%), corpus callosum hypoplasia (19%), and abnormal cerebrum (14%).

Of 22 individuals with available MRIs for review, 10 had cerebellar atrophy (8/2, mild/severe) (Supplemental Figure 3). Two had mild generalized frontal lobe volume loss and one had mild cortical thinning.

#### In-person neurological examination

Forty-nine individuals had neurological examinations at a median age of 7.9 years (range 1.7-59.5). The motor exam was notable for muscle atrophy, mixed tone and weakness (Table 1). Half had muscle atrophy of both legs (mostly affected gastrocnemius). Thirty-six individuals (73.5%) had axial hypotonia and 51% had upper extremity hypotonia. Twenty-nine individuals (59.2%) had bilateral slight increase in tone. Twenty-nine individuals (59.2%) showed weakness. Spastic diplegia was common. Eleven individuals (22.4%) had a positive or modified Gowers’ sign. Ten individuals (20.4%) had a hyperkinetic movement disorder. Coordination abnormalities were observed in 12 individuals (24.5%), with dysmetria, truncal titubation, hand tremors, jerky movements, and ataxia. Hyperreflexia was noted, mostly in the lower extremities. In ambulatory individuals, abnormal gait patterns were predominantly ataxic and/or scissoring with diplegia.

#### In-person functional assessments

Twenty-five percent (GMFCS III) walked with an assistive device, and 30.4% (GMFCS I/II) walked independently. Over half (55.6%) could handle objects without assistance (MACS I/II). Forty percent could communicate independently in most environments (CFCS I/II). Most individuals (72.7%) were able to eat and drink safely (EDACS I/II) (Supplemental Figure 4). Over half (55.6%) had age-appropriate oral intake with no restrictions (pFIOS VI). All functional parameters were correlated (Supplemental Table 5).

#### In-person motor assessments

Fifty-six individuals with a median age of 8.2 years (range 1.7-59.5) completed gross motor assessments. The mean GMFM-66 score was 49.7 out of 100 (*SD*=10.0). Mean distance walked on the 6MWT was 456.5 meters (n=12, *SD*=81.0) which represents an average of 73.3% (*SD*=15.7) of predicted for age, sex, and height. Mean 10MWR time was 4.7 seconds (n=8, *SD*=1.5) and TUG time was 8.2 seconds (n=7, *SD*=2.8). Nearly all were able to walk fast (n=13, 76.4%) and most were able to run (n=11, 64.7%). Both stride length and cadence increased with fast paced and running conditions. Mean percent time in double support during preferred paced walking (32.8%, *SD*=9.2) was greater than norms (20.6%, *SD*=2.9)^23^. Mean RULM score on the dominant side was 26.8 out of 37 (n=16, *SD*=10.8, range 6-37). Mean of BBT was 29.3 blocks (n=18, *SD*=15.6, range=10-61), with mean z-score -5.6 (*SD*=2.6, range -10.8 to -0.5) ^24,25^. On average, 9HPT times were 53.6 seconds (*SD*=46.4, range=18-227) with mean z-score -8.4 (*SD* = 10.5, range -39.1 to 0.5)^26^. There were moderate to strong associations between GMFM-66 scores and all other ambulatory and upper limb functional assessments (Supplemental Figure 5).

### Ophthalmologic features

By caregiver report, ophthalmologic problems were noted in 84% and included optic nerve atrophy (53%), strabismus (31%), and cortical visual impairment (27%). Optic nerve atrophy was diagnosed at a median age of 4 years (*IQR*=5).

In-person ophthalmologic assessment was performed on 24 individuals, although not all individuals were able to participate in all examination due to age or neurocognitive disability. The average central visual acuity in children was 20/43 Snellen (logMAR 0.3, range 0-0.8), and 20/119 Snellen (logMAR 0.8, range 0.5-1.4) in adults. On examination using slit lamp biomicroscopy, 21% of individuals exhibited mild optic nerve atrophy and 63% of individuals had moderate to severe optic nerve atrophy. Ninety-five percent of individuals had thinned retinal nerve fiber layer (RNFL) and ganglion cell layer-inner plexiform layer (GCL-IPL) thicknesses on optical coherence tomography, consistent with optic nerve atrophy. Ninety-two percent of individuals had color vision deficits on Hardy Rand Rittler color plate testing. Thirty-eight percent had strabismus or history of strabismus, and 85% of individuals who were able to participate in stereopsis testing had impaired stereo vision. Visual fields to confrontation were constricted in 92% of individuals who were able to participate in testing. Visual evoked potentials in three individuals showed varying degrees of signal slowing and diffuseness, indicating dysfunctional signal transmission from the optic nerve to the posterior visual pathways.

When comparing ophthalmologic findings, there were significant differences between caregiver report and in-person ophthalmologic assessment (Supplemental Table 6).

### Developmental and behavioral symptoms

By caregiver report, common behavioral challenges included sleep difficulties (42%), attention-deficit/hyperactivity disorder (25%), and autism (24%). Developmental regression was reported in 27%.

In-person cognitive assessments were conducted in 25 individuals with a median age of 9.3 years (range 3.8-59.5). Individuals unable to participate in cognitive assessments included 29.8% who were non-verbal and could not respond to commands, had severe visual or motor impairment, English was not their primary language (14%), age <2.5 years (3.5%), and logistically inconvenient (8.8%). The mean composite score of General Conceptual Ability (GCA) was 61.7 (*SD*=25.6; median=52, range 30-117). Verbal abilities were in the low range (*M*=71.6, *SD*=28.1; median=65, range 30-113) but were significantly higher than the spatial score (*M*=57.8, *SD*=29.1; median=46, range 32-118), but not significantly different from the nonverbal reasoning score (*M*=66.2, SD=20.4; median=63, range 31-101) (Supplemental Figure 6). GMFM-66 was moderately correlated with all DAS-II subdomain scores; and the logMar (higher logMAR indicates poor vision acuity) was negatively correlated to the spatial score only (Supplemental Table 7). Two older individuals in ages range 40-44 and 55-59 years, both had general ability index scores of 71 (verbal comprehension score 78 and 68, and perceptual reasoning score 69 and 79).

Baseline VABS was collected on 146 individuals with a median age of 8.2 years (range 1-58.3). The mean VABS-ABC at baseline was 62.9 (*SD*=19.1). The socialization score (*M*=66.6, *SD*=21.5) was significantly higher than daily living skills (*M*=58.3, *SD*=22.1), and motor skills score (*M*=58.5, *SD*=19.8) but not different with the communication score (*M*=60.7, *SD*=22.4) (Figure 2A). VABS-ABC was strongly correlated with verbal (*r*=0.71, *p*<0.01), and moderately correlated with GCA, nonverbal reasoning, and spatial score on DAS-II (Supplemental Table 7).

**Figure 2:**
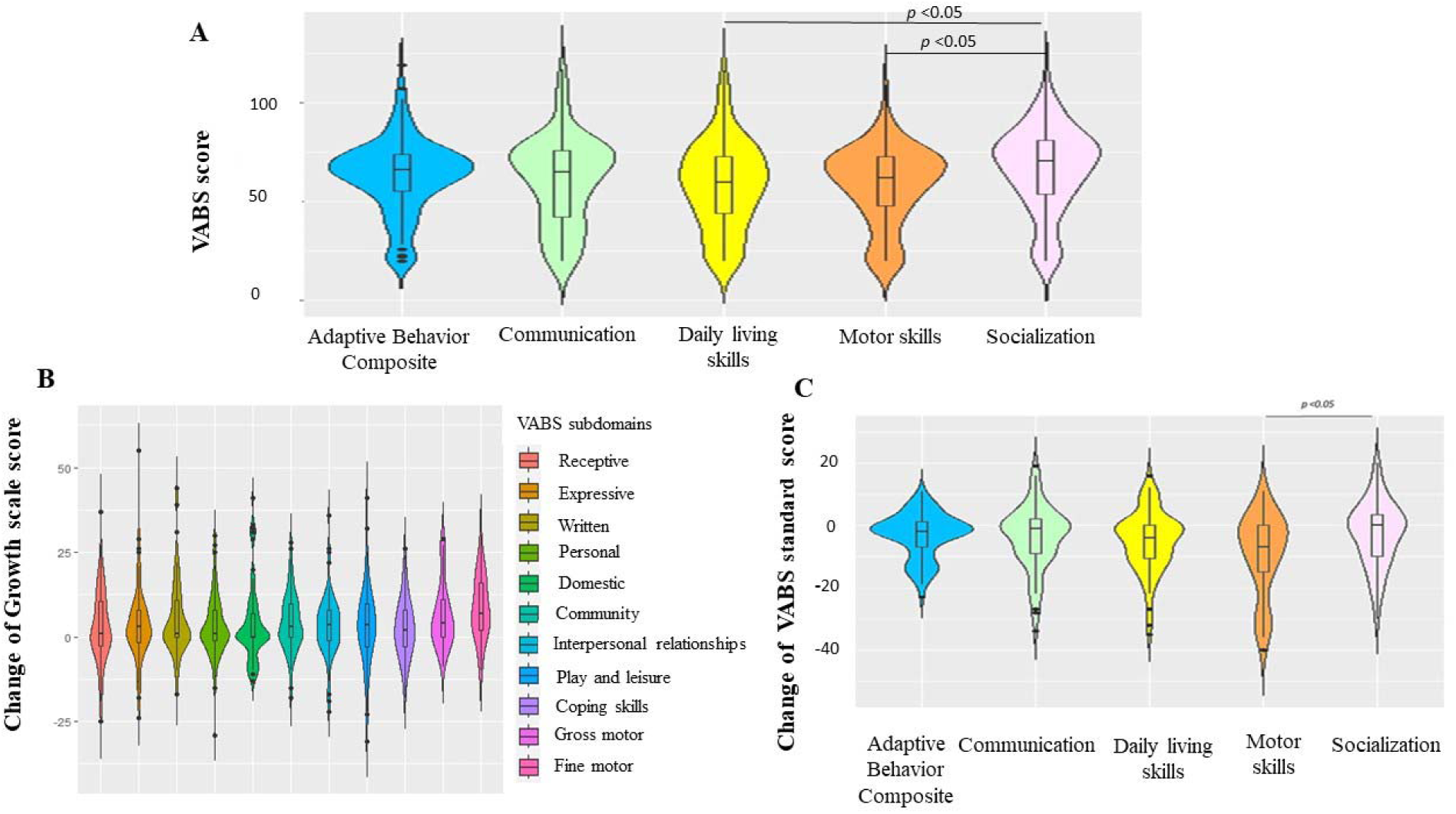
(A) Violin plots illustrate Vineland Adaptive Behavior Scales 3rd Edition (VABS) standard score of individuals with pathogenic/likely pathogenic *KIF1A* variants at baseline (n=146). Each domain standard score has a mean of 100 and a standard deviation of 15. Socialization score was significantly higher than daily living skill, and motor skill score. (B) Violin plots illustrate differences between baseline and follow-up of growth scale value in each subdomain (n=80). There was no difference between growth scale value in each subdomain. (C) Violin plots illustrate differences between baseline and follow-up of VABS standard score in adaptive behavior composite and each subdomain (n=80). The change of socialization score was significantly different from motor skill subdomain.

Follow-up VABS was collected in 80 individuals. The mean duration between follow-up was 2.5 years (*SD*=0.7). The mean Growth Scale Value (GSV) scores of each subdomain ranged from 2.6-8.5. The highest median GSV score was the fine motor subdomain (median=7, range - 10 to 30) and lowest was the domestic subdomain (median=0, range -13 to 41) (Figure 2B). The mean change in VABS-ABC was -3.1 (*SD*=7.3, range -23 to 11). Follow-up VABS-ABCs were significantly lower in individuals when compared to their baseline across all subdomains (p<0.05) (Supplement Table 8). Fifty-nine percent of the individuals’ VABS-ABC decreased at follow up and 10% of individuals had decreased VABS-ABC ≥1SD. The change in socialization standard score (*M*=-2.7, *SD*=10.4) was significantly less than the change in motor skill (*M*=-9.3, *SD*=13.5) and daily living skills (*M*=-5.1, *SD*=9.9); *p*<0.05, but no difference with the change of communication standard score (*M*=-3.9, *SD*=10.3); *p*>0.05 (Figure 2C).

### Correlation between clinical and genetic factors with VABS scores

#### Clinical phenotypes and VABS scores

VABS-ABCs were negatively correlated with seizures and abnormal EEG and positively correlated with ability to walk without assistance (Figure 3A). We performed PCA with phenotypes and VABS scores for all individuals. Baseline VABS-ABCs have high loading on the first principal component (PC1), age of first medical visit and hypotonia have high loading on PC2 (Supplemental Figure 7). Individuals with abnormal EEG/seizures have lower VABS-ABCs across all ages of assessment (Figure 3B). The change in VABS-ABC of individual is highly dependent on the age at first assessment. About half of individuals with baseline assessment at age younger than 6.9 had substantial decrease in scores at the follow-up assessment. Most of the individuals with baseline assessment at age older than 6.9 do not have substantial decrease in the follow-up assessment (Figure 3C). Furthermore, individuals with abnormal EEG/seizures usually had abnormal neuroimaging and optic nerve atrophy (Supplement Figure 8).

**Figure 3:**
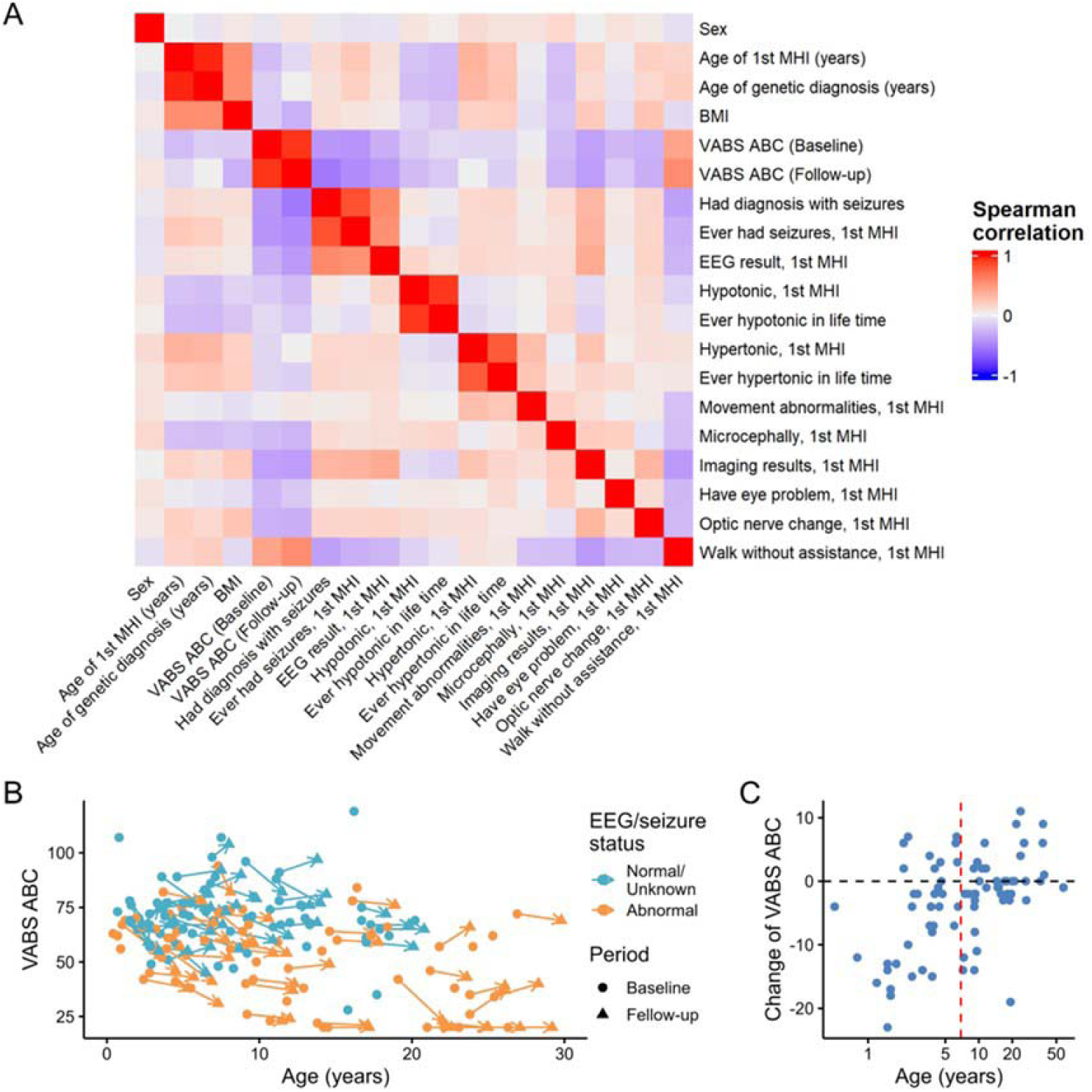
Overview of clinical phenotypes and VABS-ABC. (A) Correlation among baseline and clinical phenotypes. The order is determined by hierarchical clustering. There are 3 major blocks of phenotypes with moderate to high correlation within each block: (a) age and of 1st medical visit and diagnosis, (b) VABS-ABC, and (c) EEG, seizure, and structural disorders. (B) VABS-ABC over age of assessment. Each point represents an assessment, the points representing baseline and follow-up assessments of the same individuals are connected by an arrow. Some individuals only have one assessment. Individuals are colored by EEG or seizure status. In general, individuals with abnormal EEG or seizure have lower VABS-ABC. (C) Change of VABS-ABC between two assessment is associated with the age of 1st assessment. About half of individuals with baseline assessment at age younger than 6.9 years had substantial decrease of scores in the follow-up assessment. Most of the individuals with baseline assessment at age older than 6.9 do not have substantial decrease in the follow-up assessment. The dashed red line represents the median age (6.9 years).

#### Association of clinical and genetic factors with VABS-ABCs

Given the dependence of change in VABS-ABCs on the age at baseline assessment, in the following analysis we focused on two metrics: the change of VABS-ABCs for individuals with baseline assessment before age of 6.9 (“change of VABS-ABC”, N=79) and the final VABS-ABCs assessed after age of 6.9 (“final VABS-ABC”, N=144). We found abnormal EEG/seizures was significantly associated with lower final VABS-ABCs (beta=-20, *p*<0.001) and marginally associated with larger decrease in VABS-ABCs (beta=-6, *p*=0.03) (Figure 4A, Supplemental Table 9-10). Abnormal MRI results were significantly associated with lower final VABS-ABCs (beta=-22, *p*<0.001) (Figure 4B, Supplement Table 9-10) but not the change in VABS-ABCs (*p*=0.75).

**Figure 4:**
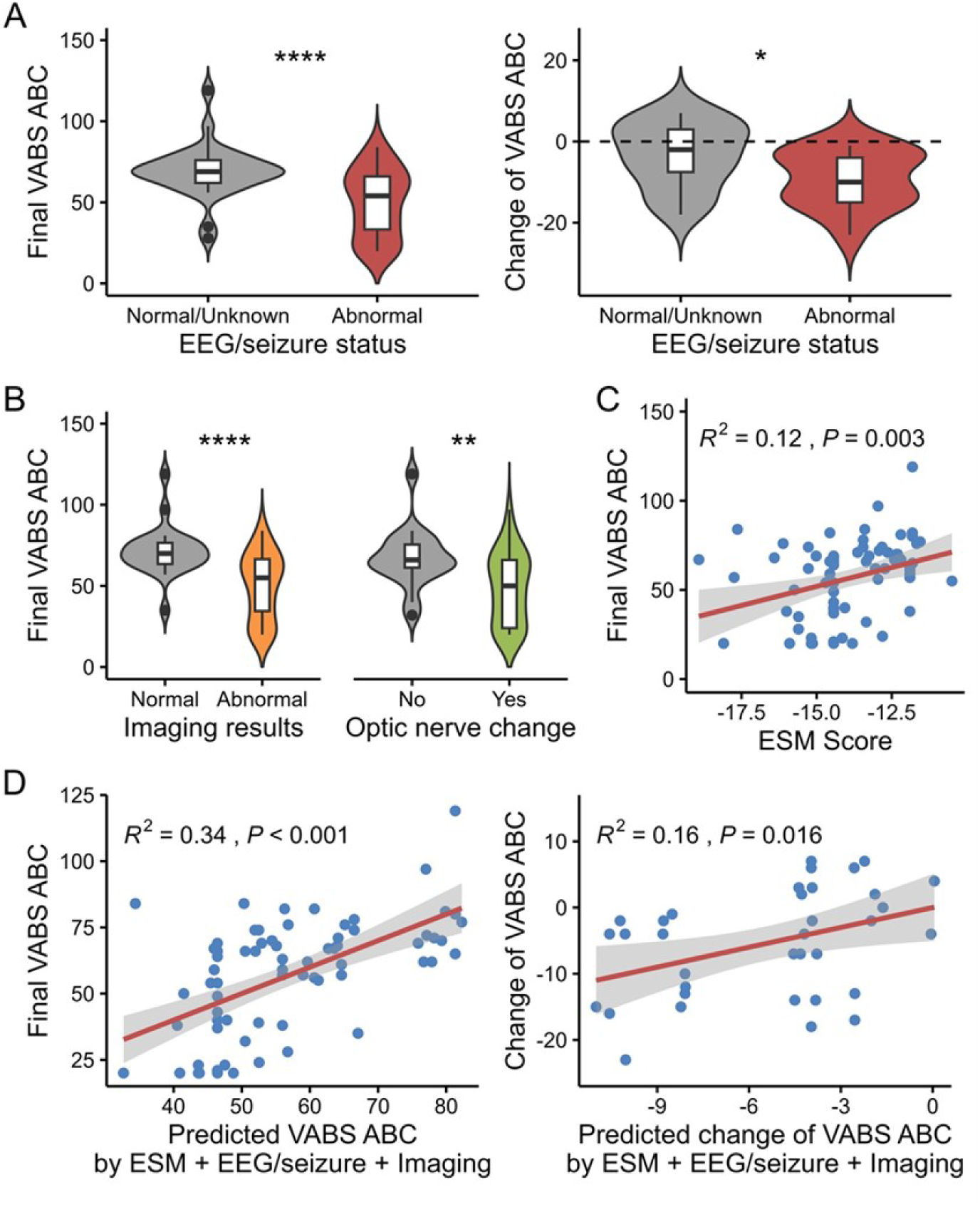
Association of clinical and genetics factors on Vineland ABC score. (A) Association of EEG or seizure status on final and change of VABS-ABC. Individuals with abnormal EEG or seizure have both lower final and change of VABS-ABC. (B) Association of neuroimaging results and optic nerve change on final VABS-ABC. Individuals with abnormal neuroimaging results or altered optic nerve showed lower final VABS-ABC than the reference group. (C) There is a significant positive correlation between ESM score and final VABS-ABC. (D) Strong positive correlations were found between predicted and observed final VABS-ABC, as well as predicted and observed change of VABS-ABC. Significance: * P < 0.05, ** P < 0.01, *** P < 0.001, **** P < 0.0001.

Some genetic variants were present in multiple individuals in the cohort, with R316W, R254W, and P305L as the three most common variants. Individuals with P305L had a trend toward higher final VABS-ABCs and smaller change in VABS-ABCs compared to the cohort (Supplemental Figure 9). Although all the missense variants observed in the cohort are pathogenic, their severity is not uniform, and the distribution of ESM score ranges -16 to -10 (Supplemental Figure 10). There was a significant positive correlation between ESM and final VABS-ABCs score (R^2^ = 0.12, *p*=0.003, Figure 4C). However, no significant correlation was observed between ESM score and abnormal EEG/seizures or neuroimaging results (Supplemental Figures 11-12). We performed multivariate linear regression of final and change in VABS-ABCs using EEGs/seizures, neuroimaging results, and ESM scores (Supplement Table 11-12). These three factors explain about 34% of the variance in the final VABS-ABCs (*p*<0.001) and about 16% of the change in VABS-ABCs (*p*=0.016) (Figure 4D).

## Discussion

The most common clinical characteristics in our cohort were hypotonia, spasticity, difficulty with coordination, ataxia, seizures, neuropathy, optic nerve atrophy, cerebellar atrophy, and cognitive impairment, similar to the previous studies^1,27–31^. This cohort which spans the age of 6 months to 58 years with an average age at follow-up of 9.1 years shows that some individuals are young and earlier in the disease course and are gaining developmental skills while others are losing skills and show signs of neurodegeneration. This heterogeneity across alleles and through the life course makes it challenging to provide accurate prognostic information to families.

We report a high frequency of seizures and/or abnormal EEGs, similar to previous studies^29–31^. The caregiver report of epilepsy is reliable and was verified by medical record review and/or review of clinical EEGs. These results confirm that *KIF1A* should be considered an epilepsy gene and that baseline EEG is warranted in newly diagnosed patients.

Although ambulatory individuals had a range of gait speeds, gait assessments revealed deficits in endurance and balance. Quantitative gait analysis showed that individuals have longer double support time as is often observed in individuals with cerebellar ataxia or unstable gait^32,33^. Future assessments of gait may benefit from formal gait analyses, pose estimation studies or other kinematic video analysis to quantify motor coordination. The combination of spasticity, coordination challenges, visual defect, and risk of osteopenia^34^, many individuals walk quickly but fall and frequently fracture bones leading to prolonged immobilization which often leads to loss of functionality. A balance of safety and independence can be achieved with appropriate adaptive equipment.

Individuals also have fine motor difficulties with lower z-scores on both BBT and NHPT. Moreover, NHPT are severely impaired with a wide range of z-score. The NHPT requires precise fine motor skills, coordination, and good vision to perform the task, so these impairments contribute to poor performance. Awareness of these limitations is important in planning individualized educational plans.

Ophthalmologic problems are pervasive, and optic nerve atrophy has likely been underdiagnosed. With in-person ophthalmologic assessment, we report much higher frequency of optic nerve atrophy, color vision deficits, depth perception defect, and visual fields impairment than caregivers reported. Since individuals with KAND may be cognitively impaired, it may be difficult for them to describe their visual symptoms, especially if they have never had normal vision. We recommend regular assessments with a neuro-ophthalmologist to assess for optic nerve atrophy and cataracts that we have observed in some older individuals.

From previous reports, cognitive function in individuals with KAND ranges from normal to profound intellectual disability^28,30,31,35^. We used standard cognitive and adaptive function assessments to quantify the cognitive profile. However, individuals with KAND have significant comorbidities of visual, fine motor, and coordination impairments which may variably interfere with the cognitive assessment. We observed very low mean GCA scores with a wide range from superior to profound cognitive impairment. The verbal scores were higher than spatial score, and this differential may be due to the confounding visual and motor impairments. Additionally, verbal scores were not correlated with logMAR. Verbal scores thus likely provide a more accurate assessment of cognitive function in this population. Furthermore, overall adaptive function (VABS-ABC) had the highest correlation with verbal ability. Socialization skills were a relative strength and better than daily living skills, which are likely limited by visual, motor, and coordination impairments.

We are beginning to characterize the longitudinal disease course over a 2.5-year period of VABS follow-up. From GSV, individuals demonstrated a wide range of change in adaptive function. VABS standard scores declined over time with a greater decline in the daily living skills and motor domain compared with the socialization domain. Individuals with KAND acquired some new adaptive functions over time; however, often at a slower rate (especially motoric function) compared to norms. Abnormal EEG/seizure was significantly associated with change of VABS-ABC and lower final VABS-ABC. Seizures and side effects of antiseizure medications can affect overall cognitive, adaptive function and learning^36,37^. Furthermore, we found that individuals with abnormal EEG/seizures usually had abnormal imaging and optic nerve atrophy which may reflect the more severe spectrum of disease. Whether abnormal electrical activity in the brain is leading to limitations in adaptive skills or simply a biomarker of underlying severity of neuronal dysfunction will benefit from longitudinal studies, ideally of presymptomatic individuals. We observed greater declines in adaptive function in the younger group and this may represent the more severe clinical spectrum which presents earlier clinically or is associated with seizures that affect the developing brain. Early detection and good seizure control may improve adaptive function, however, there was no single antiseizure medication associated with good seizure control across the cohort.

ESM is a protein language model with predictions highly correlated with variant effect across genes ^38^. Our cohort had a wide range of ESM scores, which may explain the broad spectrum of clinical phenotype. We found a significant positive correlation between ESM score and final VABS-ABC, suggesting the severity of functional impact of KIF1A pathogenic variants is associated with clinical severity in individuals, and that ESM scores may help in predicting the adaptive function for novel variants.

KAND is associated with a wide range of clinical phenotypes manifesting mainly with neurological, cognitive, and ophthalmologic manifestations along a continuum and that varies over the life course for a complex neurodevelopmental and neurodegenerative condition. Detailed longitudinal assessments, ideally annually, will help to better understand trajectories for different alleles and prepare for future clinical trials. Clinical trial designs will likely benefit from having each individual serve as their own control and by using novel outcome measures that can account for the complex confounds of performance on standardized measures due to the multiple impairments associated with KAND.

## Conclusion

The most common characteristics of KAND are hypotonia, spasticity, coordination difficulties, ataxia, seizures, peripheral neuropathy, cerebellar atrophy, visual impairment with optic nerve atrophy, and cognitive impairment. Most individuals had gross motor difficulties, and safe ambulation allows for independence and exploration. Optic nerve atrophy may be underdiagnosed, and adaptations for visual impairment may support safety and more effective education. Cognitive function varies widely, and social skills and communication are a strength. Adaptive function compared to norms declines over time and is associated with age at first assessment and abnormal EEG/seizures. ESM predictions of genotype severity may help in predicting the adaptive function. By better understanding the strengths and limitations and disease course, we strive to improve outcomes for individuals with KAND.

## Data Availability

All data produced in the present study are available upon reasonable request to the authors

## Acknowledgements

We acknowledge the contribution of all the individuals and their families who participated in this study and KIF1A.org. We extend our appreciation to Dr. Lia Boyle, Dr. Scott Brodie, Dr. Alban C. Ziegler, Candace Cameron, Jerome Doerger, Rebecca Hernan, Joanne Carroll, Michael Zuccaro, Emily Callejo, Kyle Zreibe, David Uher, Alison Garber, Ryan Cohen, Tina (Xinzhu) Yang, Sneha Sharma, Akina Sanyang, Gursimran Grewal, Jay Patel, Andrew Manahan, Tongzhuo Chang, Arianna DeCaro, Brianna Zavilowitz, Sarah Huang, Tiffany Wu, Lindsey Weathersbee, Yunzhe Hu, and Celia D’Amato.

