## Supplemental data for "Heterogeneity of comprehensive clinical phenotype and longitudinal adaptive function and correlation with computational predictions of severity of missense genotypes in KIF1A-associated neurological disorder"

**Supplemental Table**

**Supplemental Table 1:** Online and in-person clinical assessment

| Assessment | Detail |
| --- | --- |
| Online assessment | |
| Medical history | - Medical interview with a physician or genetic counselor via phone with a medical record review.  - Follow-up clinical data were collected via online questionnaires completed by caregivers with an interval of at least 12 months between longitudinal data collection timepoints. |
| Adaptive function   - Vineland Adaptive Behavior Scales 3rd Edition (VABS) | - Adaptive Behavior Composite (ABC), communication, daily living skills (DLS), socialization, and motor skills (age 0-9 years) standard score were used in the analyses.  - Each standard score has a mean of 100 and a standard deviation of 15  - For longitudinal adaptive function, growth scale values (GSV) were used to compare change relative to previous performance and change of VABS standard scores were used to compare rate of change relative to a normative sample. |
| Brain imaging  - Magnetic resonance imaging (MRI) | - From each session a scan was selected out of all available that best met the following criteria: T1-weighted, smallest slice thickness, non-contrast enhanced and acquired in or reconstructed to the sagittal plane.  - Images were reoriented and registered using 6 degrees of freedom to a template using a linear transformation. This provided a standard reference space and orientation to evaluate longitudinal changes.  - Gross anatomical findings were noted qualitatively including cerebellar appearance, cortical thinning and ventricular volume and assigned to a three-point qualitative scale (absent, mild, severe). |
| In-person clinical assessment | |
| Neurological assessment | In-person neurological assessment consisted of mental status, cranial nerves, motor, reflex, coordination, and gait assessments. |
| Ophthalmic assessment | In-person ophthalmic assessment consisted of a standard ophthalmic examination including best corrected visual acuity, color testing with Hardy-Rand-Rittler (HRR) pseudoisochromatic screening color plates, visual field testing to confrontation, extraocular motility examination, stereopsis testing, slit lamp biomicroscopy, dilated fundoscopy and optical coherence tomography (OCT). In individuals able to report visual acuity, measurements were collected in Snellen acuity and converted to the log of the minimum angle of resolution (logMAR) for statistical analysis. Allen optotypes were used for nonverbal participants. Humphrey visual fields were attempted when patients had capacity. Not all individuals were able to participate in all sections of the standard examination, owing to neurocognitive ability or age limitations. OCT imaging was obtained using swept-source OCT on the Cirrus 6000 device (Carl Zeiss Meditec, Inc., Dublin, CA, USA). Each OCT image and automated segmentations were reviewed. Pattern-reversal visual evoked potentials (VEP) were performed as per published International Society for Clinical Electrophysiology of Vision (ISCEV) protocols^30^. Briefly, electrodes were placed on the patients’ head in predetermined locations dictated by the ISCEV protocol. Signals were then measured as the individual focused on a pattern-reversal grid of 1^o^ and 0.25^o^ checkered boxes, or through flash VEP if the participant was unable to focus on a central target. |
| Cognitive assessment | - Cognitive assessment in individuals with significant visual and motor impairments posed potential confounds, and thus the assessment was adapted for best performance including providing more time on motor tasks, using grip-support pencil, adjusting the distance and angle of the material for supporting visual challenges.  - The General Conceptual Ability (GCA), verbal, nonverbal, and spatial cluster score from DAS-II were obtained, as well as the composite score of General Ability Index (GAI), verbal comprehension, and perceptual reasoning composite score from WAIS-IV were used in the analyses.  - Each score has a mean of 100 and a standard deviation of 15 |
| Gross motor | - Gross Motor Functional Measure assess gross motor function across 5 dimensions (lying and rolling, sitting, creeping, and kneeling, standing, and walking, running, and jumping)  - The GMFM-66 score has a maximum of 100% with higher scores indicating better function. |
| Upper extremities assessment | - The Revised Upper Limb Module assess upper limb (shoulder, mid-level elbow, wrist, and hand) function  - The Box and Blocks Test counted the number of blocks that individuals could move over a partition, one at a time, in a 60-second time period.  - The 9 Hole Peg Test, individuals were instructed to place 9 pegs each in their own hole one by one, and remove them one by one, and move them each back to their starting well as quickly as possible. Each hand was tested independently. |
| Ambulatory assessment | - The 10 Meter Walk Run measured the time in second, an individual uses to walk or run for 10 meters as quickly and safely as possible.  - The 6-Minute Walk Test measured the maximum distance in meters an individual could walk in six minutes over a 25m course.  - The Timed Up and Go measured the time in seconds an individual used to stand up from a chair, walk three meters, turn around, and return to sitting. |

**Supplemental Table 2:** Available ESM-1b score of KIF1A variants in this study.

| **cDNA notation** | **P notation** | **Inheritance** | **Final review pathogenicity** | **Variant type** | **ESM score** |
| --- | --- | --- | --- | --- | --- |
| c.1031C>T | p.T344M | De novo | Likely pathogenic | missense | -16.201 |
| c.1451G>A | p.G484D | De novo | Likely pathogenic | missense | -15.016 |
| c.160T>G | p.Y54D | Paternal | Likely pathogenic | missense | -17.636 |
| c.173C>T | p.S58L | De novo | Pathogenic | missense | -12.925 |
| c.254C>A | p.A85D | De novo | Pathogenic | missense | -17.748 |
| c.265T>G | p.Y89D | De novo | Likely pathogenic | missense | -16.117 |
| c.290G>C | p.G97A | De novo | Likely pathogenic | missense | -12.727 |
| c.296C>A | p.T99K | De novo | Likely pathogenic | missense | -16.778 |
| c.296C>T | p.T99M | Not tested | Pathogenic | missense | -15.178 |
| c.296C>T | p.T99M | De novo | Pathogenic | missense | -15.178 |
| c.304G>A | p.G102S | De novo | Pathogenic | missense | -13.046 |
| c.308A>C | p.K103T | De novo | Likely pathogenic | missense | -14.148 |
| c.31C>T | p.R11W | Maternal | Likely pathogenic | missense | -15.605 |
| c.32G>A | p.R11Q | Not tested | Pathogenic | missense | -13.452 |
| c.32G>A | p.R11Q | De novo | Pathogenic | missense | -13.452 |
| c.350G>T | p.G117V | De novo | Pathogenic | missense | -16.409 |
| c.377T>C | p.L126P | De novo | Likely pathogenic | missense | -19.406 |
| c.37C>T | p.R13C | Not tested | Pathogenic | missense | -13.29 |
| c.37C>T | p.R13C | Do novo | Pathogenic | missense | -13.29 |
| c.38G>A | p.R13H | Not tested | Pathogenic | missense | -15.276 |
| c.38G>A | p.R13H | De novo | Pathogenic | missense | -15.276 |
| c.38G>A | p.R13H | Not tested | Likely pathogenic | missense | -15.276 |
| c.442G>A | p.E148K | Not tested | Likely pathogenic | missense | -14.699 |
| c.443A>T | p.E148V | Paternal mosaic parent | Likely pathogenic | missense | -15.138 |
| c.452G>A | p.C151Y | Not tested | Likely pathogenic | missense | -14.801 |
| c.463C>T | p.R155C | Not tested | Pathogenic | missense | -12.246 |
| c.467A>T | p.D156V | De novo | Pathogenic | missense | -13.638 |
| c.469C>T | p.L157F | De novo | Likely pathogenic | missense | -10.5 |
| c.470T>A | p.L157H | Not tested | Likely pathogenic | missense | -13.813 |
| c.506G>A | p.R169K | De novo | Pathogenic | missense | -12.343 |
| c.507G>T | p.R169S | Paternal mosaic parent | Likely pathogenic | missense | -13.361 |
| c.512A>C | p.H171P | De novo | Likely pathogenic | missense | -18.92 |
| c.556G>T | p.V186F | De novo | Pathogenic | missense | -15.747 |
| c.575T>G | p.I192S | De novo | Likely pathogenic | missense | -15.438 |
| c.596G>A | p.G199E | Not tested | Pathogenic | missense | -12.816 |
| c.596G>A | p.G199E | De novo | Pathogenic | missense | -12.816 |
| c.596G>A | p.G199E | De novo | Likely pathogenic | missense | -12.816 |
| c.596G>A | p.G199R | De novo | Pathogenic | missense | -10.771 |
| c.604G>C | p.A202P | De novo | Pathogenic | missense | -16.003 |
| c.609G>C | p.R203S | De novo | Pathogenic | missense | -14.264 |
| c.617C>T | p.A206V | De novo | Likely pathogenic | missense | -14.785 |
| c.629T>C | p.M210T | Paternal | Pathogenic | missense | -11.557 |
| c.631A>C | p.N211H | Not maternal | Likely pathogenic | missense | -12.309 |
| c.641G>A | p.S214N | De novo | Likely pathogenic | missense | -14.064 |
| c.643A>C | p.S215R | Not tested | Pathogenic | missense | -12.811 |
| c.646C>T | p.R216C | Not tested | Pathogenic | missense | -12.413 |
| c.646C>T | p.R216C | De novo | Pathogenic | missense | -12.413 |
| c.647G>A | p.R216H | De novo | Pathogenic | missense | -13.351 |
| c.647G>A | p.R216H | Not tested | Pathogenic | missense | -13.351 |
| c.650C>A | p.S217Y | De novo | Pathogenic | missense | -18.097 |
| c.739G>A | p.V247M | De novo | Likely pathogenic | missense | -11.721 |
| c.742G>A | p.D248N | De novo | Likely pathogenic | missense | -11.161 |
| c.743A>G | p.D248G | De novo | Pathogenic | missense | -12.198 |
| c.746T>C | p.L249P | Not tested | Pathogenic | missense | -13.445 |
| c.751G>A | p.G251R | De novo | Likely pathogenic | missense | -9.167 |
| c.757G>A | p.E253K | De novo | Pathogenic | missense | -11.817 |
| c.760C>T | p.R254W | De novo | Pathogenic | missense | -11.9 |
| c.760C>T | p.R254W | Not maternal | Pathogenic | missense | -11.9 |
| c.760C>T | p.R254W | Not tested | Pathogenic | missense | -11.9 |
| c.760C>T | p.R254W | Paternal mosaic parent | Pathogenic | missense | -11.9 |
| c.761G>A | p.R254Q | De novo | Pathogenic | missense | -12.959 |
| c.761G>C | p.R254P | Maternal mosaic parent | Pathogenic | missense | -12.767 |
| c.773C>T | p.T258M | De novo | Pathogenic | missense | -11.815 |
| c.773C>T | p.T258M | Not tested | Pathogenic | missense | -11.815 |
| c.799G>C | p.E267Q | De novo | Likely pathogenic | missense | -15.19 |
| c.815A>G | p.N272S | Not tested | Pathogenic | missense | -13.131 |
| c.815A>G | p.N272S | De novo | Pathogenic | missense | -13.131 |
| c.816C>G | p.N272K | De novo | Likely pathogenic | missense | -14.567 |
| c.821C>T | p.S274L | Not tested | Pathogenic | missense | -13.4 |
| c.821C>T | p.S274L | De novo | Pathogenic | missense | -13.4 |
| c.824T>C | p.L275P | De novo | Likely pathogenic | missense | -12.816 |
| c.833T>C | p.L278P | De novo | Likely pathogenic | missense | -14.066 |
| c.835G>C | p.G279R | De novo | Pathogenic | missense | -12.057 |
| c.836G>A | p.G279D | De novo | Pathogenic | missense | -14.99 |
| c.839A>G | p.K280R | De novo | Likely pathogenic | missense | -11.675 |
| c.83T>C | p.I28T | De novo | Likely pathogenic | missense | -13.26 |
| c.914C>T | p.P305L | De novo | Pathogenic | missense | -14.562 |
| c.914C>T | p.P305L | Not tested | Pathogenic | missense | -14.562 |
| c.917A>G | p.Y306C | De novo | Likely pathogenic | missense | -13.218 |
| c.919C>G | p.R307G | Not tested | Likely pathogenic | missense | -15.251 |
| c.920G>A | p.R307Q | Not tested | Pathogenic | missense | -14.434 |
| c.920G>A | p.R307Q | De novo | Pathogenic | missense | -14.434 |
| c.920G>C | p.R307P | De novo | Pathogenic | missense | -15.902 |
| c.925T>C | p.S309P | Not tested | Likely pathogenic | missense | -13.564 |
| c.934A>C | p.T312P | De novo | Likely pathogenic | missense | -15.145 |
| c.938G>C | p.W313S | Not tested | Likely pathogenic | missense | -12.824 |
| c.941T>C | p.L314P | De novo | Likely pathogenic | missense | -15.091 |
| c.946C>T | p.R316W | De novo | Pathogenic | missense | -14.436 |
| c.946C>T | p.R316W | Not tested | Pathogenic | missense | -14.436 |
| c.946C>T | p.R316W | Paternal mosaic parent | Pathogenic | missense | -14.436 |
| c.947G>A | p.R316Q | De novo | Pathogenic | missense | -12.516 |
| c.947G>A | p.R316Q | Not tested | Pathogenic | missense | -12.516 |
| c.976G>C | p.A326P | De novo | Likely pathogenic | missense | -12.633 |

**Supplemental Table 3:** Other medical history from caregiver in individuals with *KIF1A* variants

| Clinical characteristics | Percentage |
| --- | --- |
| Measurement^a^  Weight (n=115)   - Underweight - Normal - Obesity   Height (n=136)   - Short stature - Normal - Tall stature | 11.3  67.8  20.9  16.2  83.1  0.7 |
| Ear, Nose, and Throat (ENT) problems (n=167)   - History of otitis media - Pressure equalization (PE) tubes - Hearing problem | 37.7  9.0  2.4 |
| Gastrointestinal problems (n=167)   - Constipation - Gastroesophageal reflux disease - Swallowing problem - Irritable bowel syndrome | 40.1  33.5  24.0  5.4 |
| Urogenital problems (n=167)   - Urinary incontinence (> 5-year-old) - History of urinary tract infection - Hydronephrosis - Hypospadias - Congenital kidney anomalies | 40.1  22.1  2.4  2.1  1.8 |
| Orthopedics problems (n=167)   - History of broken bone - Scoliosis | 25.7  18.0 |
| Dermatologic problems (n=167)   - Eczema - Hemangioma | 19.2  1.8 |
| Respiratory problems (n=167)   - History of pneumonia - Asthma/reactive airway disease | 12.0  7.8 |
| Cardiovascular problems (n=167)   - Congenital heart disease | 2.4 |
| Endocrine problems (n=167)   - Growth hormone deficiency - Abnormal thyroid function test | 3.6  2.4 |
| Developmental, mood and behavioral problems (n=165)   - Developmental regression - Attention-deficit/hyperactivity disorder - Autism spectrum disorder - Anxiety - Obsessive-compulsive disorder - Depression - Sleep problems - Self-injurious behavior | 27.3  24.8  23.6  19.4  7.3  3.6  45.5  11.5 |
| Perinatal history (n=156) | |
| Pregnancy complication (%)   - Pre-eclampsia - Multiple birth pregnancy - Gestational diabetes - Polyhydramnios | 7.1  3.8  3.2  1.3 |
| Gestational age (day); mean±SD | 39.1±1.9 |
| Birth weight (Z score); mean±SD | -0.1±1.0 |
| Birth length (Z score); mean±SD | 0.3±1.1 |
| Birth head circumference (Z score); mean±SD | -0.2±1.1 |
| Apgar score; mean±SD | 1min: 8.4±1.5, 5min: 9.3±0.8 |
| Hospital stay (day); median (IQR) | Median 2(1) |
| NICU stay (%) | 7.7 |
| Perinatal complication (%)   - Preterm birth - Small for gestational age - Respiratory distress - Hypoglycemia - Meconium aspiration - Neonatal seizure | 7.1  7.1  7.1  4.5  2.6  0.6 |
| Neonatal problems (%)   - Feeding difficulty/poor sucking - Hyperbilirubinemia - Irritable/inconsolable/colicky - Floppy infant - Lethargic - Hypertonic | 29.5  26.9  25.6  19.9  17.9  9.0 |

^a^ Measurement definition^40,41^

- Underweight: <2 years old; weight-for-length ≤z-score 2, 2–20 years old; BMI z-score <-1.6, ≥20 years old; BMI <18.5 kg/m2
- Obese: <2 years old; weight-for-length ≥z-score 2, 2–20 years old; BMI z-score ≥1.6, ≥20 years old; BMI ≥30 kg/m2
- Short stature: height-for-age; <z-score –2
- Tall stature: height-for-age; >z-score –2

**Supplemental Table 4:** Common neurological medication

| Medication | Percentage |
| --- | --- |
| Antiepileptic medications  Type of medication   - Levetiracetam - Lamotrigine - Clobazam - Valproic acid - Oxcarbazepine - Topiramate   Number of mediations used   - Single medication - Two medications - Combine 3-4 medications | 55.1  26.5  20.4  14.3  10.2  6.1  57.1  26.5  16.4 |
| Treatment for spasticity   - Botox injections - Baclofen | 34  17 |

**Supplemental Table 5:** Correlation between age and functional classification

|  | 1 | 2 | 3 | 4 | 5 | 6 |
| --- | --- | --- | --- | --- | --- | --- |
| 1. Age |  |  |  |  |  |  |
| 2. GMFCS | -0.076 |  |  |  |  |  |
| 3. MACS | -0.363 | .614** |  |  |  |  |
| 4. CFCS | -.600** | .410* | .750** |  |  |  |
| 5. EDAS | -0.113 | .711** | .787** | .480* |  |  |
| 6. pFOIS | 0.227 | -.589** | -.687** | -.498** | -.848** |  |

Note: GMFCS = Gross Motor Function Classification System, MACS = Manual Ability Classification System, CFCS = Communication Function Classification System, EDACS = Eating and Drinking Ability Classification System, pFIOS = Pediatric Feeding Oral Intake Scale

* indicates p < 0.05, **indicates p < 0.01

**Supplemental Table 6:** Number and difference between ophthalmic characteristic in caregiver report and in-person assessment

| Ophthalmic characteristic | Caregiver report | In-person assessment | *P* value |
| --- | --- | --- | --- |
| Color vision deficits | 0 | 11/12 | 0.004 |
| Visual field defect | 0 | 11/12 | 0.004 |
| Depth perception defects | 1/10 | 10/10 | 0.032 |
| Optic nerve atrophy | 10/22 | 20/22 | 0.001 |

**Supplemental Table 7:** Correlation between age, Vineland, Differential Ability Scales-II, Gross Motor Function Measure (GMFM-66), LogMAR

| Variables | 1 | 2 | 3 | 4 | 5 | 6 | 7 | 8 | 9 | 10 | 11 |
| --- | --- | --- | --- | --- | --- | --- | --- | --- | --- | --- | --- |
| Age |  |  |  |  |  |  |  |  |  |  |  |
| 1. Vineland: ABC | -0.149 |  |  |  |  |  |  |  |  |  |  |
| 2. Vineland: Com | -0.064 | .925** |  |  |  |  |  |  |  |  |  |
| 3. Vineland: DLS | -0.254 | .920** | .792** |  |  |  |  |  |  |  |  |
| 4. Vineland: Socialization | -0.159 | .924** | .836** | .784** |  |  |  |  |  |  |  |
| 5. DAS-II: CGA | 0.177 | .675** | .650** | .654** | 0.456 |  |  |  |  |  |  |
| 6. DAS-II: Verbal | 0.15 | .705** | .725** | .637** | .516* | .927** |  |  |  |  |  |
| 7. DAS-II: NVR | 0.043 | .664** | .608** | .664** | .480* | .922** | .841** |  |  |  |  |
| 8. DAS-II: Spatial | 0.149 | .591** | .543* | .606** | 0.378 | .947** | .793** | .820** |  |  |  |
| 9. GMFM-66 | 0.173 | .539** | .532** | .433** | .441** | .609** | .486* | .570** | .655** |  |  |
| 10. LogMAR | 0.266 | -0.337 | -0.342 | -0.44 | -0.135 | -0.468 | -0.25 | -0.343 | -.613* | -.677** |  |

Note: ABC = Adaptive Behavior Composite, Com = Communication, DLS = Daily living skill, GCA = General Conceptual Ability, NVR = Nonverbal reasoning, GMFM-66 = Gross Motor Function Measure, LogMAR = Logarithm of the Minimum Angle of Resolution

* indicates p < 0.05, **indicates p < 0.01

**Supplemental Table 8:** Pair t test comparing Vineland-3 standard score in adaptive behavior composite and each subdomain (n=80)

|  | Baseline | | Follow-up | | *t*(79) | *p* value |
| --- | --- | --- | --- | --- | --- | --- |
|  | *M* | *SD* | *M* | *SD* |  |  |
| Adaptive behavior composite standard score | 62.3 | 20.8 | 58.9 | 20.9 | 3.7 | <0.001 |
| Communication standard score | 60.0 | 23.9 | 56.1 | 24.8 | 3.2 | <0.01 |
| Daily living skills standard score | 58.6 | 24.0 | 53.3 | 23.1 | 4.5 | <0.001 |
| Socialization standard score | 66.0 | 23.2 | 62.9 | 23.7 | 2.3 | <0.05 |
| Motor skills standard score | 60.8 | 17.7 | 51.5 | 20.9 | 4.2 | <0.001 |

**Supplemental Table 9:** Univariate linear regression of clinical phenotypes and final Vineland ABC grouped by age.

| **Variable** | **Age ≤ 6.9 years** | | | | **Age > 6.9 years** | | | |
| --- | --- | --- | --- | --- | --- | --- | --- | --- |
|  | **N** | **Mean (SD)** | **Beta (SE)** | **P** | **N** | **Mean (SD)** | **Beta (SE)** | **P** |
| Sex |  |  |  |  |  |  |  |  |
| Male | 38 | 66.11 ± 15.74 | Reference |  | 39 | 56.05 ± 20.07 | Reference |  |
| Female | 30 | 63.50 ± 14.57 | -2.61 (3.72) | 0.486 | 36 | 58.22 ± 22.80 | 2.17 (4.95) | 0.662 |
| BMI | 68 |  | -1.21 (0.54) | **0.032** | 64 |  | -0.14 (0.44) | 0.754 |
| EEG/seizure status |  |  |  |  |  |  |  |  |
| Normal/Unknown | 41 | 70.46 ± 14.84 | Reference |  | 29 | 69.10 ± 16.27 | Reference |  |
| Abnormal | 27 | 56.59 ± 11.59 | -13.87 (3.38) | **<0.001** | 46 | 49.52 ± 20.74 | -19.58 (4.54) | **<0.001** |
| Hypotonic |  |  |  |  |  |  |  |  |
| No | 6 | 72.83 ± 20.06 | Reference |  | 14 | 65.21 ± 23.00 | Reference |  |
| Yes | 62 | 64.19 ± 14.61 | -8.64 (6.45) | 0.185 | 59 | 55.34 ± 20.83 | -9.88 (6.31) | 0.122 |
| Hypertonic |  |  |  |  |  |  |  |  |
| No | 25 | 63.76 ± 15.75 | Reference |  | 6 | 63.50 ± 22.03 | Reference |  |
| Yes | 39 | 65.56 ± 15.74 | 1.80 (4.03) | 0.656 | 67 | 57.31 ± 21.10 | -6.19 (9.02) | 0.495 |
| Botox procedure |  |  |  |  |  |  |  |  |
| No | 40 | 65.12 ± 14.14 | Reference |  | 35 | 63.26 ± 21.67 | Reference |  |
| Yes | 5 | 69.80 ± 21.48 | 4.67 (7.10) | 0.514 | 22 | 54.45 ± 20.15 | -8.80 (5.74) | 0.131 |
| Movement abnormalities |  |  |  |  |  |  |  |  |
| No | 34 | 68.65 ± 18.49 | Reference |  | 35 | 54.20 ± 23.87 | Reference |  |
| Yes | 33 | 61.06 ± 9.97 | -7.59 (3.64) | **0.041** | 37 | 61.03 ± 18.05 | 6.83 (4.97) | 0.174 |
| Neuropathy |  |  |  |  |  |  |  |  |
| No | 27 | 68.15 ± 15.28 | Reference |  | 18 | 69.39 ± 17.78 | Reference |  |
| Yes | 8 | 58.62 ± 23.84 | -9.52 (7.03) | 0.184 | 30 | 56.57 ± 21.19 | -12.82 (5.96) | **0.037** |
| Microcephaly |  |  |  |  |  |  |  |  |
| No | 50 | 68.34 ± 15.20 | Reference |  | 64 | 58.64 ± 21.11 | Reference |  |
| Yes | 12 | 52.25 ± 10.95 | -16.09 (4.67) | **0.001** | 8 | 45.00 ± 18.60 | -13.64 (7.83) | 0.086 |
| Imaging results |  |  |  |  |  |  |  |  |
| Normal | 35 | 65.23 ± 12.36 | Reference |  | 23 | 71.61 ± 15.37 | Reference |  |
| Abnormal | 28 | 62.82 ± 17.08 | -2.41 (3.71) | 0.519 | 47 | 49.98 ± 20.37 | -21.63 (4.81) | **<0.001** |
| Have eye problem |  |  |  |  |  |  |  |  |
| No | 6 | 74.17 ± 17.27 | Reference |  | 12 | 71.67 ± 8.12 | Reference |  |
| Yes | 61 | 64.07 ± 14.94 | -10.10 (6.47) | 0.123 | 62 | 54.13 ± 22.06 | -17.54 (6.48) | **0.009** |
| Optic nerve change |  |  |  |  |  |  |  |  |
| No | 28 | 63.82 ± 12.41 | Reference |  | 18 | 67.17 ± 18.47 | Reference |  |
| Yes | 28 | 62.39 ± 16.86 | -1.43 (3.96) | 0.719 | 37 | 48.78 ± 21.94 | -18.38 (6.00) | **0.003** |
| Walk without assistance |  |  |  |  |  |  |  |  |
| No | 33 | 58.45 ± 13.68 | Reference |  | 33 | 48.76 ± 19.34 | Reference |  |
| Yes | 20 | 74.00 ± 12.91 | 15.55 (3.80) | **<0.001** | 29 | 67.10 ± 19.24 | 18.35 (4.91) | **<0.001** |

**Supplemental Table 10:** Univariate linear regression of clinical phenotypes and change of Vineland ABC grouped by age.

| **Variable** | **Age ≤ 6.9 years** | | | | **Age > 6.9 years** | | | |
| --- | --- | --- | --- | --- | --- | --- | --- | --- |
|  | **N** | **Mean (SD)** | **Beta (SE)** | **P** | **N** | **Mean (SD)** | **Beta (SE)** | **P** |
| Sex |  |  |  |  |  |  |  |  |
| Male | 20 | -4.70 ± 8.68 | Reference |  | 22 | -3.05 ± 6.64 | Reference |  |
| Female | 16 | -6.50 ± 7.28 | -1.80 (2.71) | 0.511 | 21 | 0.76 ± 4.75 | 3.81 (1.77) | 0.037 |
| BMI | 36 |  | -0.44 (0.20) | **0.038** | 43 |  | 0.17 (0.16) | 0.310 |
| EEG/seizure status |  |  |  |  |  |  |  |  |
| Normal/Unknown | 23 | -3.35 ± 7.92 | Reference |  | 13 | -1.54 ± 6.15 | Reference |  |
| Abnormal | 13 | -9.31 ± 6.94 | -5.96 (2.63) | **0.030** | 30 | -1.03 ± 6.10 | 0.51 (2.03) | 0.805 |
| Hypotonic |  |  |  |  |  |  |  |  |
| No | 4 | 1.25 ± 6.55 | Reference |  | 7 | -2.00 ± 5.32 | Reference |  |
| Yes | 32 | -6.34 ± 7.87 | -7.59 (4.12) | 0.074 | 36 | -1.03 ± 6.23 | 0.97 (2.52) | 0.702 |
| Hypertonic |  |  |  |  |  |  |  |  |
| No | 14 | -7.14 ± 8.42 | Reference |  | 4 | -0.25 ± 8.73 | Reference |  |
| Yes | 22 | -4.45 ± 7.77 | 2.69 (2.74) | 0.334 | 38 | -1.32 ± 5.93 | -1.07 (3.25) | 0.745 |
| Botox procedure |  |  |  |  |  |  |  |  |
| No | 18 | -7.72 ± 7.76 | Reference |  | 20 | -0.70 ± 6.40 | Reference |  |
| Yes | 4 | -1.25 ± 5.38 | 6.47 (4.12) | 0.132 | 15 | -2.67 ± 6.80 | -1.97 (2.24) | 0.387 |
| Movement abnormalities |  |  |  |  |  |  |  |  |
| No | 18 | -5.67 ± 8.85 | Reference |  | 20 | -0.80 ± 7.04 | Reference |  |
| Yes | 18 | -5.33 ± 7.36 | 0.33 (2.71) | 0.903 | 22 | -1.59 ± 5.28 | -0.79 (1.91) | 0.681 |
| Neuropathy |  |  |  |  |  |  |  |  |
| No | 15 | -5.73 ± 8.55 | Reference |  | 7 | -8.71 ± 7.95 | Reference |  |
| Yes | 3 | -8.00 ± 8.72 | -2.27 (5.42) | 0.681 | 20 | 1.80 ± 4.25 | 10.51 (2.36) | **<0.001** |
| Microcephaly |  |  |  |  |  |  |  |  |
| No | 27 | -4.48 ± 8.36 | Reference |  | 38 | -1.21 ± 6.39 | Reference |  |
| Yes | 8 | -9.38 ± 6.28 | -4.89 (3.21) | 0.136 | 4 | -1.25 ± 2.75 | -0.04 (3.25) | 0.990 |
| Imaging results |  |  |  |  |  |  |  |  |
| Normal | 20 | -6.20 ± 8.37 | Reference |  | 10 | -1.10 ± 5.15 | Reference |  |
| Abnormal | 14 | -5.29 ± 7.94 | 0.91 (2.86) | 0.751 | 30 | -1.20 ± 6.03 | -0.10 (2.13) | 0.963 |
| Have eye problem |  |  |  |  |  |  |  |  |
| No | 2 | -7.50 ± 10.61 | Reference |  | 2 | -11.50 ± 0.71 | Reference |  |
| Yes | 34 | -5.38 ± 8.04 | 2.12 (5.91) | 0.722 | 41 | -0.68 ± 5.72 | 10.82 (4.10) | **0.012** |
| Optic nerve change |  |  |  |  |  |  |  |  |
| No | 17 | -6.06 ± 7.00 | Reference |  | 9 | -1.33 ± 3.28 | Reference |  |
| Yes | 14 | -6.57 ± 8.79 | -0.51 (2.83) | 0.858 | 27 | 0.04 ± 6.25 | 1.37 (2.19) | 0.536 |
| Walk without assistance |  |  |  |  |  |  |  |  |
| No | 17 | -9.71 ± 7.03 | Reference |  | 22 | 0.41 ± 4.77 | Reference |  |
| Yes | 11 | -1.18 ± 6.21 | 8.52 (2.60) | **0.003** | 15 | -3.53 ± 8.01 | -3.94 (2.10) | 0.069 |

**Supplemental Table 11:** Multivariate linear regression of ESM, EEG/seizure status, imaging results, optic nerve change and final Vineland ABC in later years individuals.

| **Variable** | **Crude** | | **Adjusted^1^** | |
| --- | --- | --- | --- | --- |
|  | **Beta (SE)** | **P** | **Beta (SE)** | **P** |
| ESM | 3.58 (1.28) | 0.007 | 3.55 (1.30) | 0.008 |
| EEG/seizure status | -8.92 (5.86) | 0.133 | -8.61 (6.01) | 0.157 |
| Imaging results | -16.22 (6.01) | 0.009 | -16.45 (6.18) | 0.010 |

^1^ P values were adjusted for sex and age.

**Supplemental Table 12:** Multivariate linear regression of ESM, EEG/seizure status, imaging results, optic nerve change and change of Vineland ABC in early years individuals.

| **Variable** | **Crude** | | **Adjusted^1^** | |
| --- | --- | --- | --- | --- |
|  | **Beta (SE)** | **P** | **Beta (SE)** | **P** |
| ESM | -0.18 (0.98) | 0.853 | -0.26 (0.88) | 0.773 |
| EEG/seizure status | -6.24 (2.90) | 0.040 | -3.51 (2.75) | 0.213 |
| Imaging results | 1.96 (2.86) | 0.499 | 1.88 (2.56) | 0.469 |

^1^ P values were adjusted for sex and age.

**Supplemental Figures**

| Age (Year) | Number of individuals |
| --- | --- |
| [0-5] | 72 |
| (5-10] | 45 |
| (10-15] | 18 |
| (15-20] | 22 |
| (20-25] | 8 |
| (25-30] | 5 |
| (30-35] | 1 |
| (35-40] | 4 |
| (>40] | 2 |

**Supplemental Figure 1:** Histogram of the age distribution of individuals with *KIF1A* Variants at enrollment (N=177). Most of the individuals were minors <15 years old (76.3%) and 41% were less than 5 years old.

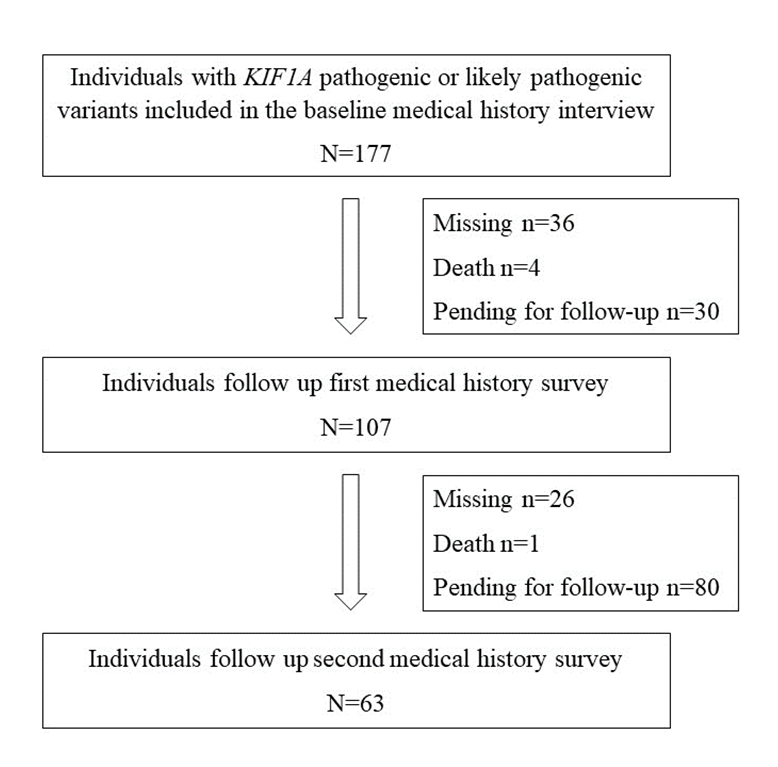

**Supplemental Figure 2:** Cohort study flow of online clinical data collection

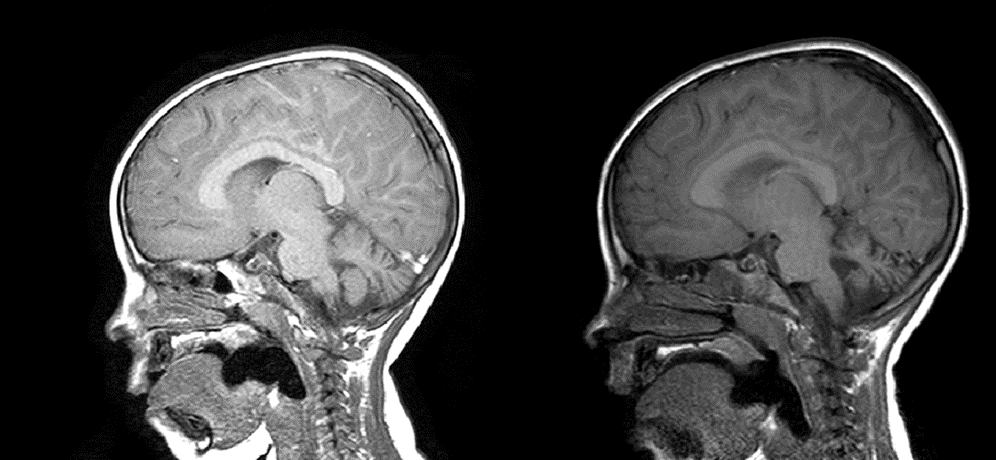

**Supplemental Figure 3:** The left panel shows a sagittal slice of an MRI of an individual at age range 0-5 years old. The right panel shows the same individual three years later. There is a reduction of overall cerebellar volume.

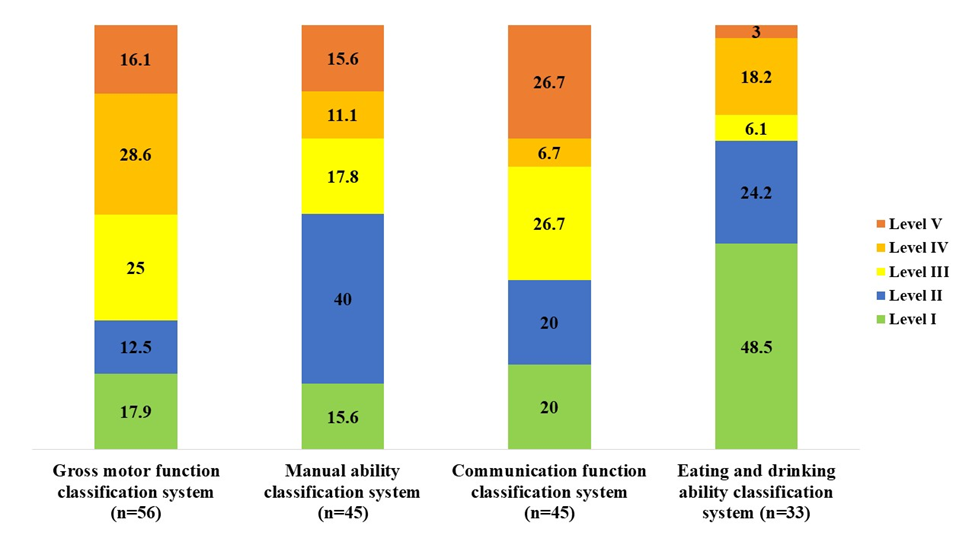

**Supplemental Figure 4:** The stacked bar chart show percentage of individual current functional classification level. The classification system provides 5 ordinal levels (I, II, III, IV, V); higher levels indicate poorer function, as levels I indicate the best function and levels V indicate the worst function.

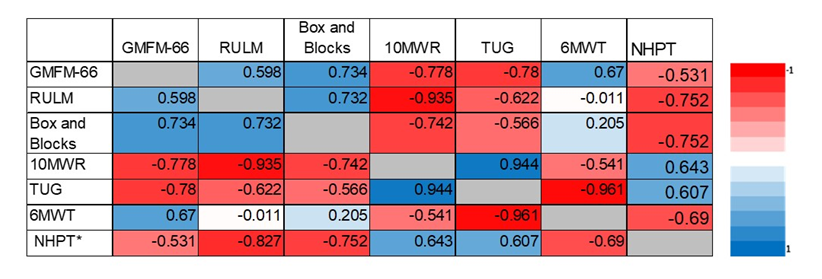

**Supplemental Figure 5:** Heat map depicting strength of correlations between Gross Motor Function Measure (GMFM)-66 and related functional tests: Revised Upper Limb Module (RULM), Box and Blocks, 10 Meter Walk Run (10MWR), Timed up and Go (TUG), Six Minute Walk Test (6MWT), Nine Hole Peg Test (NHPT).

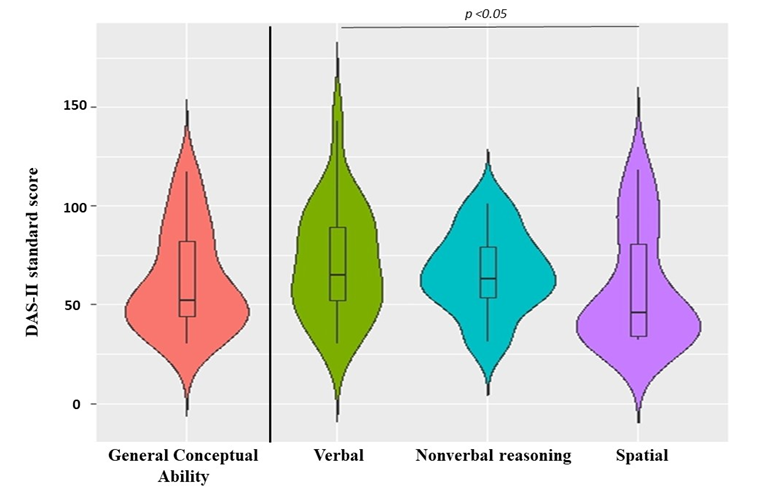

**Supplemental Figure 6:** Violin plots illustrate The Differential Ability Scales second edition (DAS-II) score of general conceptual ability and each subdomain of individuals with pathogenic/likely pathogenic KIF1A variants at baseline (n=23). Each score has a mean of 100 and a standard deviation of 15 in the general population. DAS-II was categorized to above average (score 110-119), average (score 90-109), below average (score 80-89), low (score 70-79), and very low (score ≤69). Verbal score was significantly higher than spatial score.

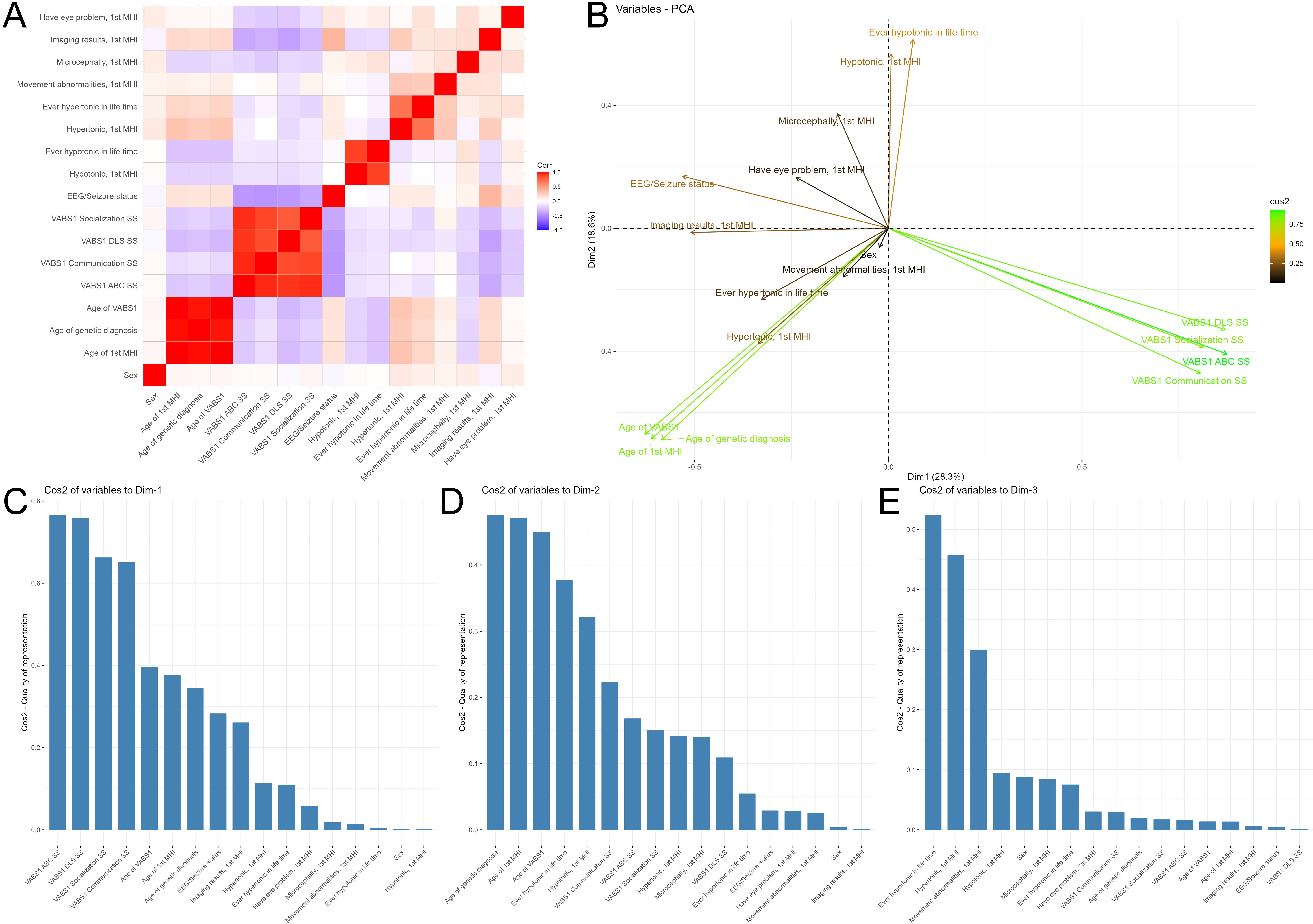

**Supplemental Figure 7:** Principal component analysis (PCA) of all individuals. (A) Heatmap of the correlation coefficient matrix for main clinical features. (B) Biplot for main clinical features, the color indicates the cos2 score. (C) Contribution of each variable on PC1. (D) Contribution of each variable on PC2. (E) Contribution of each variable on PC3.

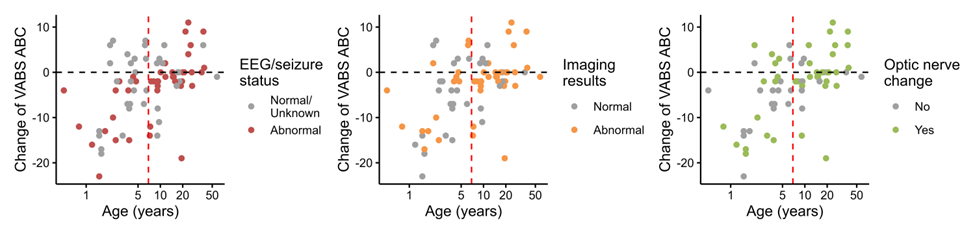

**Supplemental Figure 8:** Correlation between age and change of Vineland ABC score grouped by clinical phenotypes.

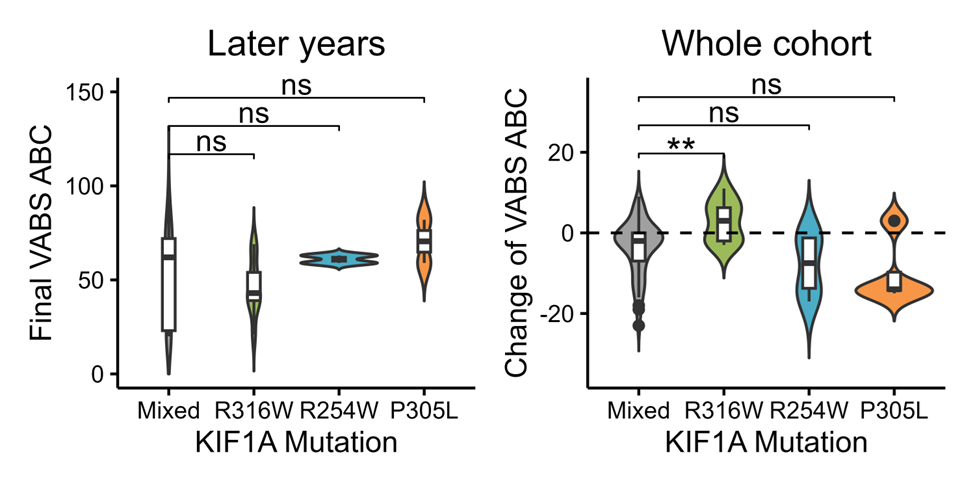

**Supplemental Figure 9:** Correlation of the top3 highly recurrent mutations and final and change of Vineland ABC score. Significance: * P < 0.05, ** P < 0.01, *** P < 0.001, **** P < 0.0001.

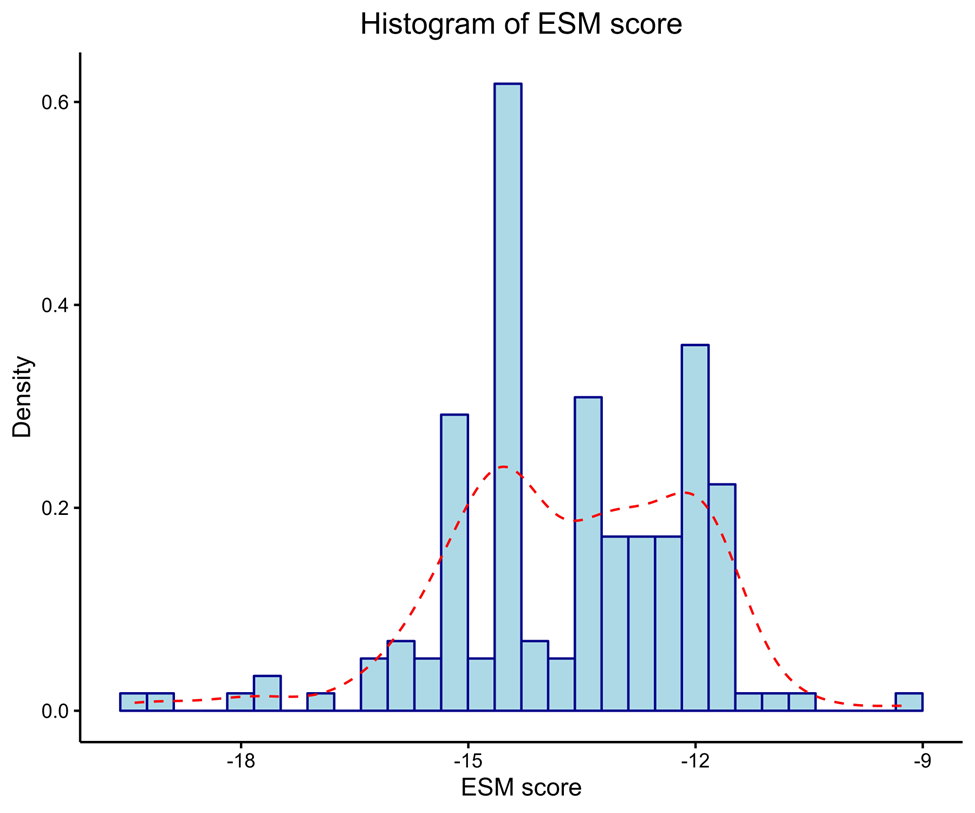

**Supplemental Figure 10:** Distribution of ESM score of KIF1A variants in this study.

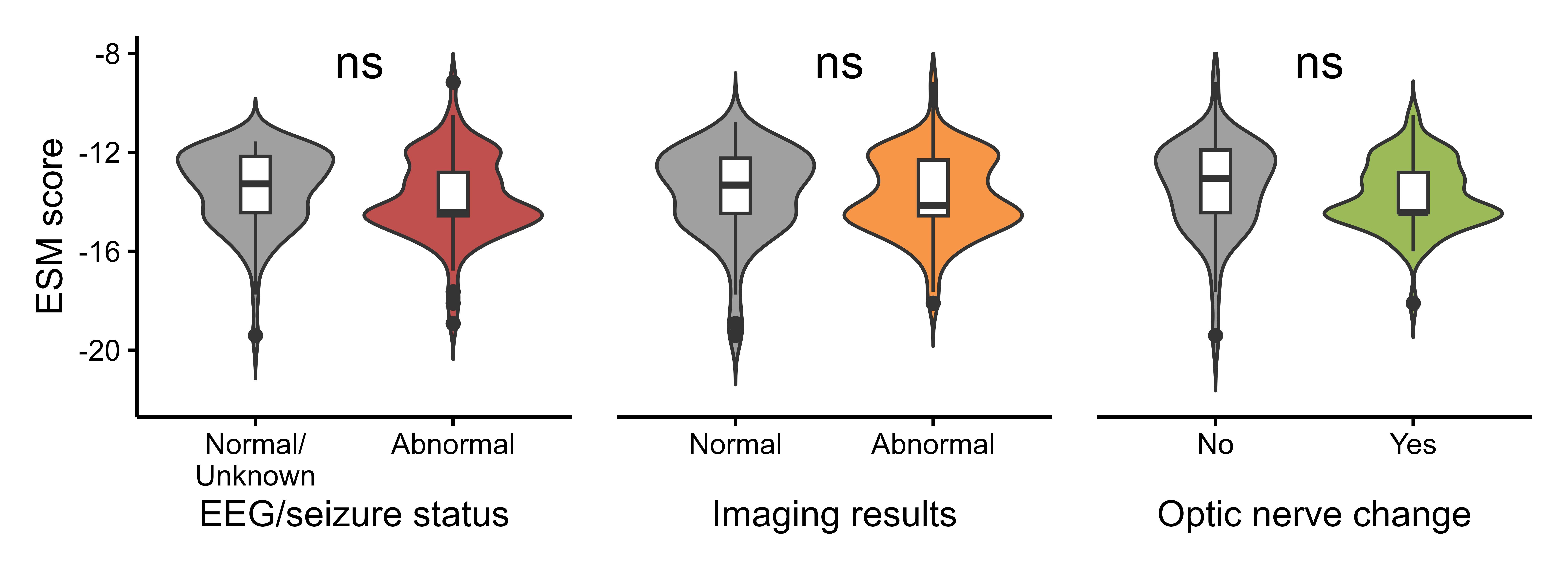

**Supplemental Figure 11:** There was no difference of ESM score between EEG or seizure status, imaging results or optic nerve change.

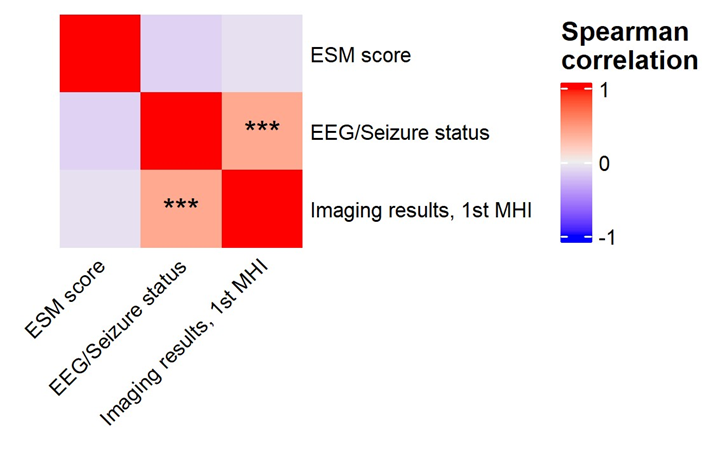

**Supplemental Figure 12:** Correlation among ESM score, EEG or seizure status, and imaging results. Significance: * P < 0.05, ** P < 0.01, *** P < 0.001, **** P < 0.0001.
